# Circadian syndrome and incident non-neoplastic digestive disease in middle-aged and older Chinese adults: a prospective cohort study

**DOI:** 10.64898/2026.09.24.26363842

**Authors:** Lu Liu, Liqiong Zhao, Cailan Li, Yan Geng

## Abstract

**Background:** Circadian syndrome (CircS) comprises metabolic abnormalities, short sleep, and depressive symptoms, and is associated with cardiovascular, liver, and lung diseases and mortality; evidence on digestive disease is scarce. We assessed its association with incident non-neoplastic digestive disease in Chinese adults.

**Methods:** In this prospective analysis of the China Health and Retirement Longitudinal Study (CHARLS), we included 2011 baseline participants aged ≥45 years free of digestive disease, with follow-up to 2018. CircS (≥4 of 7 components) was analysed as a continuous score and as a binary exposure. Cox models estimated hazard ratios (HRs) adjusting for demographic factors, BMI, smoking, and alcohol use. We did dose–response, component, subgroup, and sensitivity analyses; hs-CRP mediation was explored.

**Findings:** 907 (19·0%) of 4 771 participants developed digestive disease over a median 84 months. Each additional CircS component raised the risk by 9·1% (HR 1·091, 95% CI 1·041–1·144, P<0·001); CircS positivity (36·1%) was associated with a 25·7% higher risk (HR 1·257, 1·084–1·456, P=0·002). Risk rose approximately linearly to about 70% higher at 5–7 components. Short sleep (HR 1·178, 1·017–1·365) and depressive symptoms (HR 1·570, 1·367–1·803) were the only independently associated components; no metabolic component was associated. Results were robust across sensitivity analyses; hs-CRP did not mediate the association.

**Interpretation:** CircS is associated with incident non-neoplastic digestive disease, driven by its circadian–emotional rather than metabolic components. Sleep and mood may be targets for prevention.

**Funding:** This research received no specific grant from any funding agency in the public, commercial, or not-for-profit sectors.

## Introduction

Digestive diseases include gastro-oesophageal reflux disease (GERD), peptic ulcer, functional gastrointestinal disorders, and inflammatory bowel disease. These conditions impose a substantial burden on health systems worldwide, particularly among older adults, and the burden is growing in China as the population ages: analyses of Global Burden of Disease 2021 data estimated that peptic ulcer disease affects roughly one in six Chinese adults and gastro-oesophageal reflux disease more than one in twenty^1,2^. Functional gastrointestinal disorders also account for a substantial share of gastroenterology outpatient visits. Beyond traditional biological risk factors, lifestyle and psychosocial factors are increasingly recognised as contributors to digestive disease.

Circadian disruption, which includes short sleep, irregular sleep patterns, and accompanying mood disturbances, may contribute to gastrointestinal disease. It can influence gastrointestinal health through changes in autonomic balance, hypothalamic–pituitary– adrenal axis activity, gastrointestinal motility, mucosal barrier function, and the gut microbiota^3,4^. Short sleep and insomnia have been linked to GERD and irritable bowel syndrome (IBS)^5,6^, and depression and anxiety predict new-onset functional dyspepsia and IBS, with a bidirectional brain–gut pathway^7,8^. However, most studies examined single factors (such as short sleep or depression) and rarely considered the joint effects of co-occurring factors.

Zimmet and colleagues proposed the circadian syndrome (CircS) in 2019^9^. It combines five metabolic components (central obesity, elevated triglycerides, reduced high-density lipoprotein cholesterol, elevated blood pressure, and impaired fasting glucose) with short sleep and depressive symptoms, reflecting the co-occurrence of metabolic, behavioural, and emotional dysregulation in modern lifestyles. CircS research has focused mainly on cardiovascular and metabolic outcomes: CircS outperforms the conventional metabolic syndrome in predicting cardiovascular disease and death^10–12^ and is associated with chronic liver disease, chronic lung disease^13,14^, cognitive decline, and functional disability^15,16^. More recently, a UK Biobank cohort of 330 925 participants linked CircS to incident GERD (HR 1·20, 95% CI 1·16–1·23); in component-wise analyses, depression (HR 1·38) and short sleep (HR 1·29) were the strongest contributors^17^. That study, however, focused on a single disease in a European population and modelled the components separately without mutual adjustment. It remains unclear whether CircS predicts overall digestive disease risk and which components drive the association. Evidence from Asian populations is especially limited.

Using the CHARLS nationwide prospective cohort, this study assessed the association of baseline CircS and its continuous component score with incident non-neoplastic digestive disease. We further examined the dose–response relationship, identified independently associated components using mutually adjusted models, assessed robustness through sensitivity and subgroup analyses, and explored potential mechanisms through mediation analysis.

## Methods

### Study design and population

The cohort frame of this study is the China Health and Retirement Longitudinal Study (CHARLS),^18^ which follows a multistage probability sample of Chinese households and residents aged 45 years or older in 28 provinces. Fieldwork began in 2011, and participants were re-interviewed in 2013, 2015, and 2018. Ethical approval was granted by Peking University’s Biomedical Ethics Review Committee (IRB00001052-11015 for the household survey; IRB00001052-11014 for blood sampling), and every participant gave written informed consent. Reporting follows the STROBE recommendations for cohort studies^19^.

At the 2011 baseline, 17 708 individuals were interviewed. We then excluded 8 682 individuals aged <45 years, with BMI <15 or >45 kg/m^2^, or with hs-CRP ≥10 mg/L (to exclude acute inflammatory states); 2 123 with self-reported physician-diagnosed digestive disease at baseline; 660 without fasting blood samples; and 1 472 with missing CircS components or covariates, or without any follow-up information (exclusion reasons overlapped; details in Figure 1). The final complete-case analysis included 4 771 participants.

**Figure 1.**
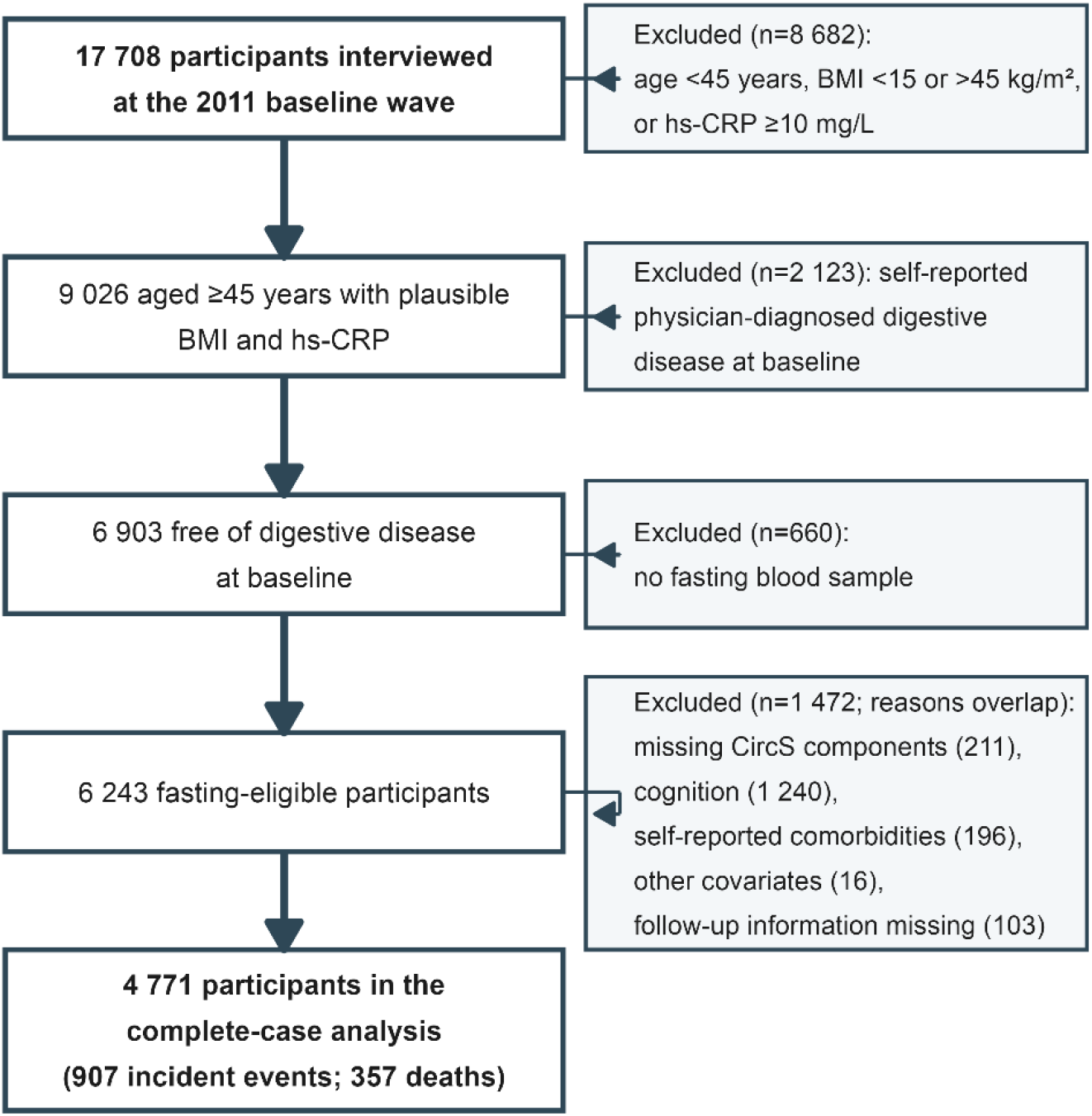
Flow diagram of participant selection.

### Exposure: circadian syndrome

CircS was defined as meeting at least four of seven criteria proposed by Zimmet and colleagues: (1) central obesity, defined as waist circumference ≥85 cm in men and ≥80 cm in women; (2) elevated triglycerides, defined as ≥150 mg/dL or self-reported use of lipid-lowering medication; (3) reduced HDL-C, defined as <40 mg/dL in men and <50 mg/dL in women; (4) elevated blood pressure, defined as systolic ≥130 mmHg or diastolic ≥85 mmHg, or self-reported use of antihypertensive medication; (5) elevated fasting glucose, defined as ≥100 mg/dL or self-reported use of glucose-lowering medication; (6) short sleep, defined as <6 h per night; and (7) depressive symptoms, defined as a score of at least 10 on the 10-item Center for Epidemiologic Studies Depression Scale (CES-D-10)^20^.

The continuous score was the number of components present (range 0–7). CircS positivity was defined as ≥4 components; ≥3 components served as an alternative cutoff in sensitivity analysis. Waist circumference, blood pressure, fasting glucose, and lipids were objectively measured at baseline, whereas sleep duration and depressive symptoms were self-reported.

### Outcome

The outcome was the first self-report of “physician-diagnosed stomach disease or other digestive disease” during follow-up (in any of the 2013, 2015, and 2018 waves). This item is independent of “cancer or malignant tumour” in the CHARLS chronic disease checklist, so digestive cancers were not included in the outcome. Event time was the interview date of the wave at which the outcome was first reported. Participants who died during follow-up were censored; the time of death was taken from the registered year and month of death when available, and was otherwise approximated by the midpoint of the fieldwork period of the wave in which death was registered (around July 2013, July 2015, and July 2018). Participants lost to follow-up were censored at their last attended wave.

### Covariates

Covariates were selected based on previous literature and causal assumptions. They included age, sex, education (primary or below, middle school, high school or above), marital status (married vs. other), residence (urban vs. rural), smoking, alcohol drinking, and BMI. We considered demographic and behavioural covariates (age, sex, education, marital status, residence, smoking, and alcohol use) as potential common causes of CircS and digestive disease^21^. We therefore adjusted for them in all adjusted models (Models 2, 3, and 4). BMI wasincluded in the primary model. It is an established risk factor for metabolic dysfunction anddigestive disease, such as reflux related to increased intra-abdominal pressure. BMI is not acomponent of CircS. Omitting BMI would leave confounding by general adiposity. Bycontrast, self-reported physician diagnoses of hypertension, diabetes, and dyslipidaemia werenot included in any model. We considered them downstream consequences of thecorresponding CircS components rather than confounders. Adjusting for them wouldintroduce overadjustment bias and block part of the total association between exposure andoutcome. Model 4 additionally included chronic kidney disease, heart disease, and stroke.These comorbidities do not overlap with the CircS definition. Cognitive function, chronicpain, hearing, vision, and hs-CRP were used for description or mechanism exploration.

### Statistical analysis

Baseline characteristics were compared between participants with and without CircS. Continuous variables are summarised as mean ± SD or median (IQR) and compared using t tests or Mann–Whitney U tests; categorical variables are summarised as n (%) and compared using chi-square tests. Standardised mean differences (SMDs) are also reported. Cox proportional hazards models were used: model 1 was unadjusted; model 2 adjusted for age, sex, education, marital status, and residence; model 3 (primary) additionally adjusted for BMI, smoking, and alcohol use; model 4 further adjusted for chronic kidney disease, heart disease, and stroke (exploratory). The exposure was modelled as a continuous score (per one-component increase) and as a binary variable (≥4 vs <4 components); HRs with 95% CIs are reported.

The proportional hazards assumption was assessed using Schoenfeld residuals. On the basis of these results, we conducted a time-stratified analysis comparing the first 36 months of follow-up with the period thereafter and tested whether the CircS–outcome association differed between the two periods. This analysis was secondary and hypothesis-generating; it was not interpreted as effect modification.

Dose–response was evaluated using restricted cubic splines with three knots at the 10th, 50th, and 90th percentiles (corresponding to 1, 3, and 5 components), together with a grouped Cox model (0, 1, 2, 3, 4, and 5–7 components) and a test for linear trend. For component analysis, two models were used, both with model 3 covariates: one included all seven components simultaneously with mutual adjustment to estimate independent associations; the other included one component at a time without adjusting for the others to estimate total associations. Subgroup analyses were stratified by sex, age (<60 vs ≥60 years), residence, BMI (<24 vs ≥24 kg/m^2^), and smoking, with interaction tests between CircS and each subgroup variable. Because 10 interaction tests were performed, subgroup differences are reported descriptively only. Kaplan–Meier event-free survival curves were generated and compared using the log-rank test.

Sensitivity analyses included: (1) excluding events within 2 years of follow-up; (2) excluding participants with baseline heart disease or stroke; (3) restricting BMI to 18·5–35 kg/m^2^; (4) a CircS cutoff of ≥3 components; (5) omitting BMI from the primary model; (6) Fine–Gray competing-risk regression with death as the competing event; (7) multiple imputation (mice package, 20 imputations; only covariates and outcome-side variables were imputed—because the exposure definition relies on fasting blood tests, which imputation cannot recover, 6 243 participants were the upper limit of the analysable sample); (8) a negative-control analysis replacing CircS with four randomly generated exposures (two continuous, two binary) in the primary model; and (9) a mediation analysis with baseline hs-CRP (log-transformed) as the mediator, using the product-of-coefficients method with 1 000 bootstrap resamples—given that hs-CRP and the exposure were measured concurrently at baseline, the mediation timeline is hypothetical and the results are mechanistic exploration only.

A two-sided α of 0·05 was used. Analyses were performed in R 4·4 (survival, rms, tableone, cmprsk, mice, survminer, and ggplot2 packages).

## Results

### Baseline characteristics

Among the 4 771 participants (mean age 58·9±8·9 years; 48·5% women), 1 724 (36·1%) were CircS positive. Over a median follow-up of 84 months, 907 (19·0%) developed digestive disease and 357 (7·5%) died. Compared with CircS-negative participants, CircS-positive participants were older, more often women, had higher BMI and hs-CRP and higher rates of self-reported hypertension, diabetes, and dyslipidaemia, but smoked and drank less (Table 1). Excluded participants were older, more often women, and less educated—an expected pattern, as cognitive test completion and follow-up participation are themselves related to age, sex, and education. However, measured metabolic indicators (fasting glucose, triglycerides, HDL-C, blood pressure) were highly similar between the two groups (all SMDs <0·13; Table S1), suggesting limited selection bias at the exposure assessment level.

**Table 1.** Baseline characteristics by CircS status (n=4 771)

| Characteristic | Overall (n=4 771) | CircS <4<br>(n=3 047) | CircS ≥4<br>(n=1 724) | P | SMD |
| --- | --- | --- | --- | --- | --- |
| Age, years | 58.9±8.9 | 58.5±9.1 | 59.5±8.6 | <0.001 | 0.112 |
| Female, n (%) | 2 313 (48.5) | 1 269 (41.6) | 1 044 (60.6) | <0.001 | 0.385 |
| BMI, kg/m<sup>2</sup> | 23.8±3.6 | 22.8±3.3 | 25.5±3.4 | <0.001 | 0.817 |
| Education: primary school or below | 3 080 (64.6) | 1 912 (62.8) | 1 168 (67.7) | 0.002 | 0.105 |
| Education: junior high school | 1 102 (23.1) | 738 (24.2) | 364 (21.1) |  |  |
| Education: senior high school or above | 589 (12.3) | 397 (13.0) | 192 (11.1) |  |  |
| Married, n (%) | 4 248 (89.0) | 2 752 (90.3) | 1 496 (86.8) | <0.001 | 0.111 |
| Urban residence, n (%) | 1 877 (39.3) | 1 085 (35.6) | 792 (45.9) | <0.001 | 0.211 |
| Smoking, n (%) | 2 000 (41.9) | 1 419 (46.6) | 581 (33.7) | <0.001 | 0.265 |
| Alcohol use, n (%) | 2 118 (44.4) | 1 468 (48.2) | 650 (37.7) | <0.001 | 0.213 |
| Self-reported dyslipidaemia | 477 (10.0) | 147 (4.8) | 330 (19.1) | <0.001 | 0.452 |
| Self-reported hypertension | 2 346 (49.2) | 1 124 (36.9) | 1 222 (70.9) | <0.001 | 0.725 |
| Self-reported diabetes | 760 (15.9) | 276 (9.1) | 484 (28.1) | <0.001 | 0.504 |
| Chronic kidney disease | 203 (4.3) | 114 (3.7) | 89 (5.2) | 0.024 | 0.069 |
| Heart disease | 492 (10.3) | 220 (7.2) | 272 (15.8) | <0.001 | 0.271 |
| Stroke | 110 (2.3) | 49 (1.6) | 61 (3.5) | <0.001 | 0.122 |
| hs-CRP, mg/L, median [IQR] | 1.0 [0.6, 1.9] | 0.9 [0.5, 1.7] | 1.3 [0.7, 2.5] | <0.001 | 0.294 |
| Cognitive score | 12.0±3.5 | 12.1±3.5 | 11.8±3.6 | 0.001 | 0.101 |
| Chronic pain | 1 335 (28.0) | 739 (24.3) | 596 (34.6) | <0.001 | 0.228 |
| Hearing: good/fair/poor | 2 274/1 945/552 | 1 509/1 218/320 | 765/727/232 | <0.001 | 0.119 |
| Vision: good/fair/poor | 1 118/3 208/445 | 790/2 022/235 | 328/1 186/210 | <0.001 | 0.206 |
| CircS component score | 3.0±1.6 | 2.0±0.9 | 4.7±0.8 | <0.001 | 3.045 |

Values are mean ± SD, median [IQR], or n (%); SMD, standardized mean difference.

### CircS and risk of digestive disease

Without adjustment, each additional CircS component was associated with a 5·1% higher risk (HR 1·051, 95% CI 1·009–1·095, P=0·017). After adjustment for demographic factors alone, this association weakened and became non-significant (HR 1·035, 0·992– 1·079, P=0·116). The binary exposure followed a similar pattern (model 1: HR 1·160, P=0·029; model 2: HR 1·106, P=0·148). In the primary model, which further adjusted for BMI, smoking, and alcohol use, risk increased by 9·1% per additional component (HR 1·091, 1·041–1·144, P<0·001), and CircS-positive participants (≥4 components) had a 25·7% higher risk (HR 1·257, 1·084–1·456, P=0·002) (Table 2). The pattern of attenuation followed by strengthening across models suggests that confounding was not unidirectional, consistent with the negative confounding by BMI described below.

**Table 2.** Association between CircS and incident digestive disease (Cox models; n=4 771, 907 events)

| **Model** | **Continuous (per +1 component),**<br>**HR (95% CI)** | **Binary ($\geq 4$ vs $<4$), HR (95% CI)** |
| --- | --- | --- |
| Model 1 (unadjusted) | 1.051 (1.009–1.095), P=0.017 | 1.160 (1.015–1.326), P=0.029 |
| Model 2 (demographics†) | 1.035 (0.992–1.079), P=0.116 | 1.106 (0.965–1.269), P=0.148 |
| Model 3 (primary‡) | 1.091 (1.041–1.144), P<0.001 | 1.257 (1.084–1.456), P=0.002 |
| Model 4 (fully adjusted§) | 1.082 (1.032–1.134), P=0.001 | 1.229 (1.059–1.425), P=0.006 |

† age, sex, education, marital status, residence; ‡ model 2 + BMI, smoking, alcohol use; § model 3 + chronic kidney disease, heart disease, stroke (exploratory).

After further adjustment for chronic kidney disease, heart disease, and stroke, the results were similar (per one-component increase HR 1·082, 1·032–1·134; binary HR 1·229, 1·059– 1·425). After removing BMI, the association was attenuated and no longer significant (per one-component increase HR 1·034, 0·991–1·079; binary HR 1·106, 0·965–1·269), indicating negative confounding by BMI (Table S6).

### Dose–response relationship

The dose–response curve was approximately linear (non-linearity P=0·76; knots at 1, 3, and 5 components; Figure 2). With the 0-component group as reference, the HRs for 1, 2, 3, 4, and 5–7 components were 1·14 (0·80–1·63), 1·15 (0·82–1·62), 1·36 (0·96–1·91), 1·42 (1·00–2·03), and 1·70 (1·18–2·44), respectively (trend P<0·001; Table S8). Point estimates rose markedly from 3 components onwards, and the risk elevation at 5–7 components was statistically significant.

**Figure 2.**
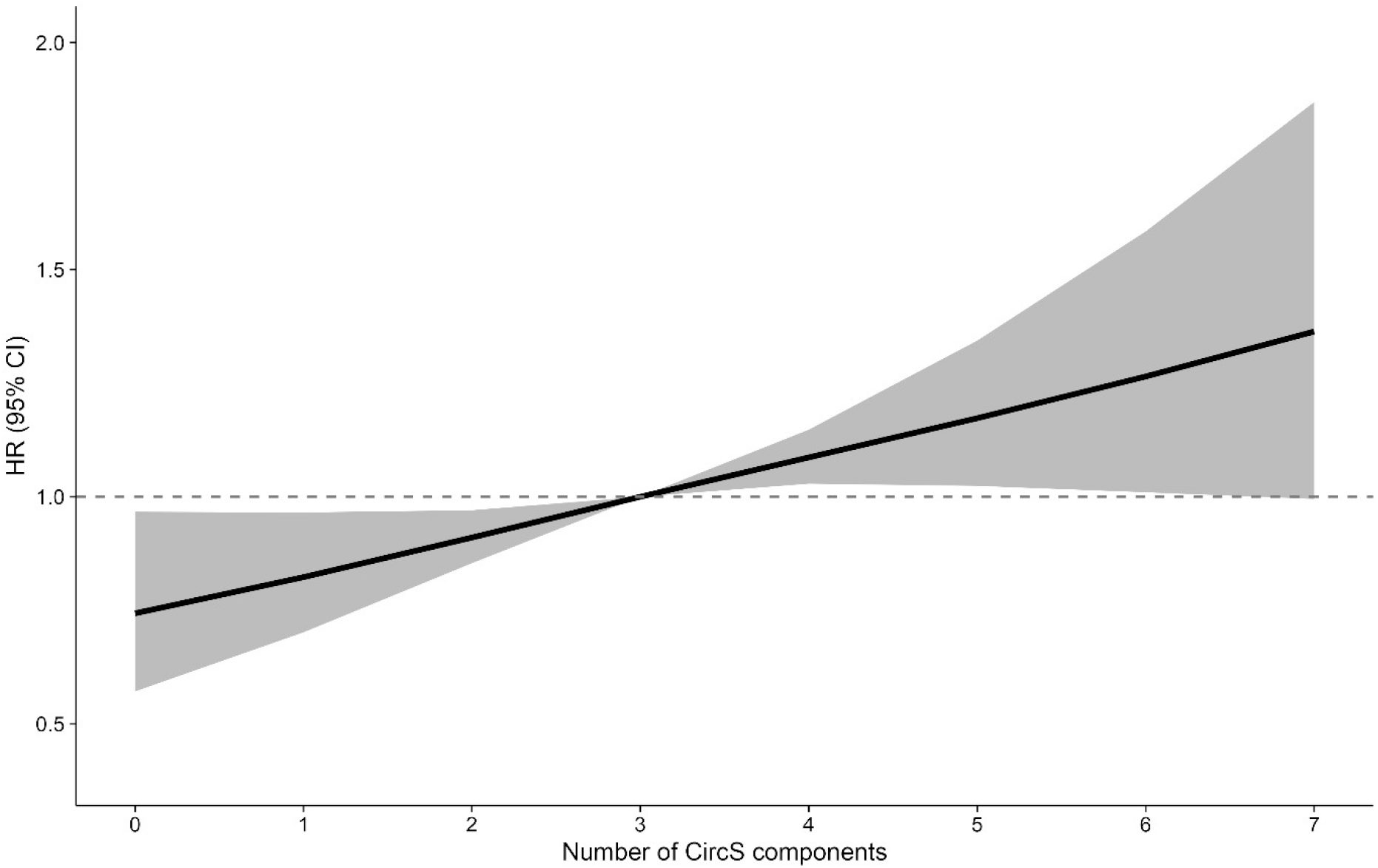
Dose–response association between the number of CircS components and incident digestive disease (restricted cubic spline with 3 knots at 1, 3, and 5 components; adjusted for model 3 covariates). The shaded area indicates the 95% CI; non-linearity P=0·76.

### Time-stratified analysis

The overall Schoenfeld residual test was not significant (P=0·21), no covariate violated the assumption (all P≥0·05), and only CircS showed borderline time variation (P=0·046) (Table S7). The time-stratified analysis (secondary, hypothesis-generating only) showed: during the first 36 months (123 events), neither the continuous exposure (HR 0·996, 0·876– 1·132) nor binary CircS (HR 1·127, 0·753–1·687) was associated with the outcome; after 36 months (4 530 participants, 784 events), each one-component increase raised the risk by 10·7% (HR 1·107, 1·053–1·165, P<0·001), and CircS positivity (≥4 components) by 27·9% (HR 1·279, 1·092–1·499, P=0·002) (Figure S1). However, the period-by-exposure interaction was not significant (continuous P=0·174; binary P=0·607) (Tables S2 and S3).

### Component-specific associations

When all 7 components were entered simultaneously (mutual adjustment), only 2 components retained independent associations: short sleep (HR 1·178, 1·017–1·365) and depressive symptoms (HR 1·570, 1·367–1·803); none of the 5 metabolic components (central obesity, triglycerides, HDL-C, blood pressure, fasting glucose) was independently associated (all P>0·40) (Table S4, Figure 3). Single-component models gave similar estimates for these 2 components (HRs 1·291 and 1·617, respectively).

**Figure 3.**
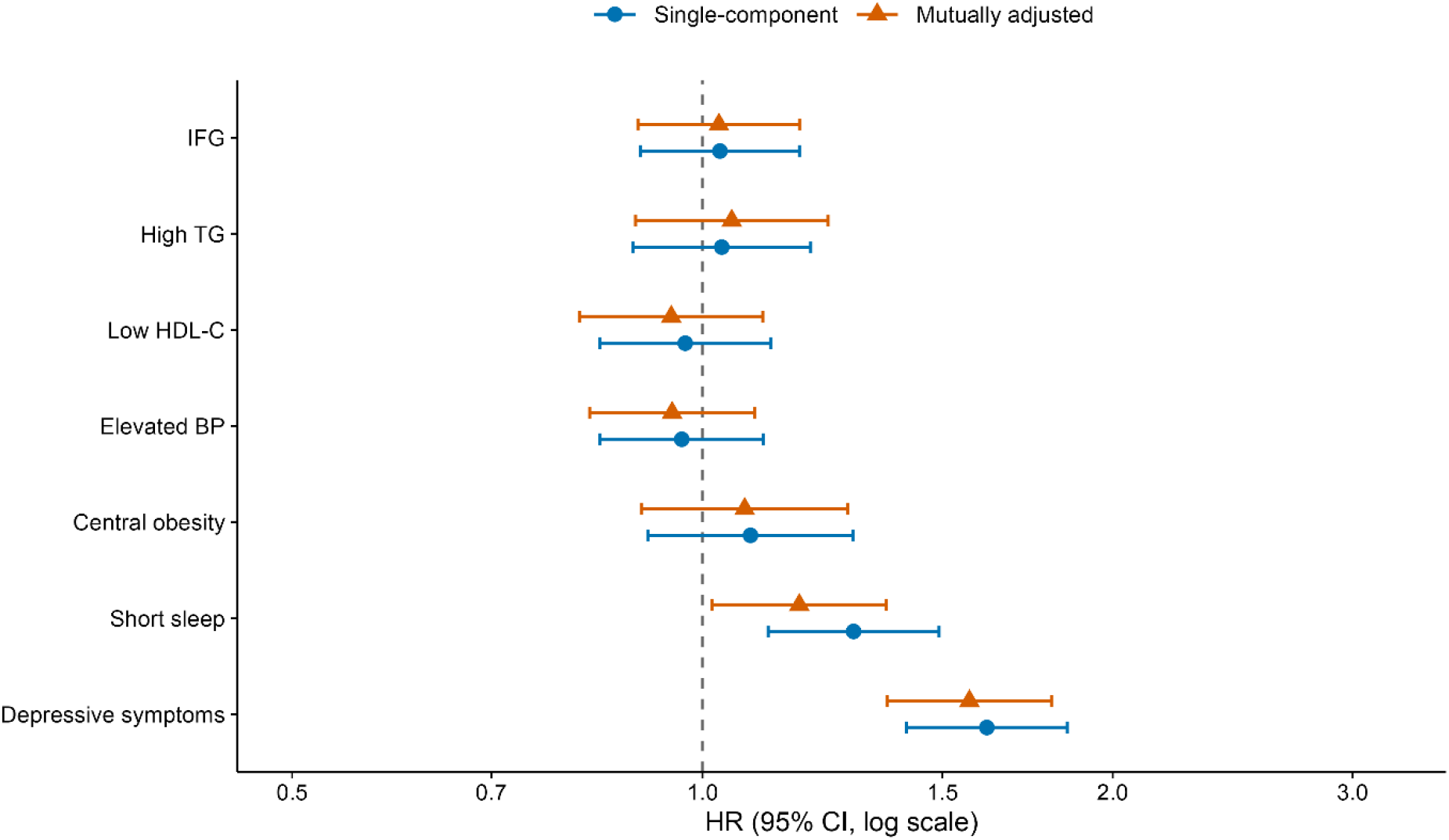
Associations of the 7 CircS components with incident digestive disease: single-component models and the mutually adjusted model (all adjusted for model 3 covariates).

### Subgroup analysis

The association was directionally consistent across subgroups, with higher point estimates in women (HR 1·440, 1·193–1·738, vs 1·015, 0·790–1·304 in men), participants younger than 60 years (1·379 vs 1·092), non-smokers (1·416 vs 1·000), and those with BMI <24 kg/m^2^ (1·315 vs 1·212). Interaction P values for binary CircS were 0·032 (sex) and 0·049 (age); however, none of the 10 interaction tests remained significant after Bonferroni correction, so these patterns are presented descriptively only (Figure 4, Table S9).

**Figure 4.**
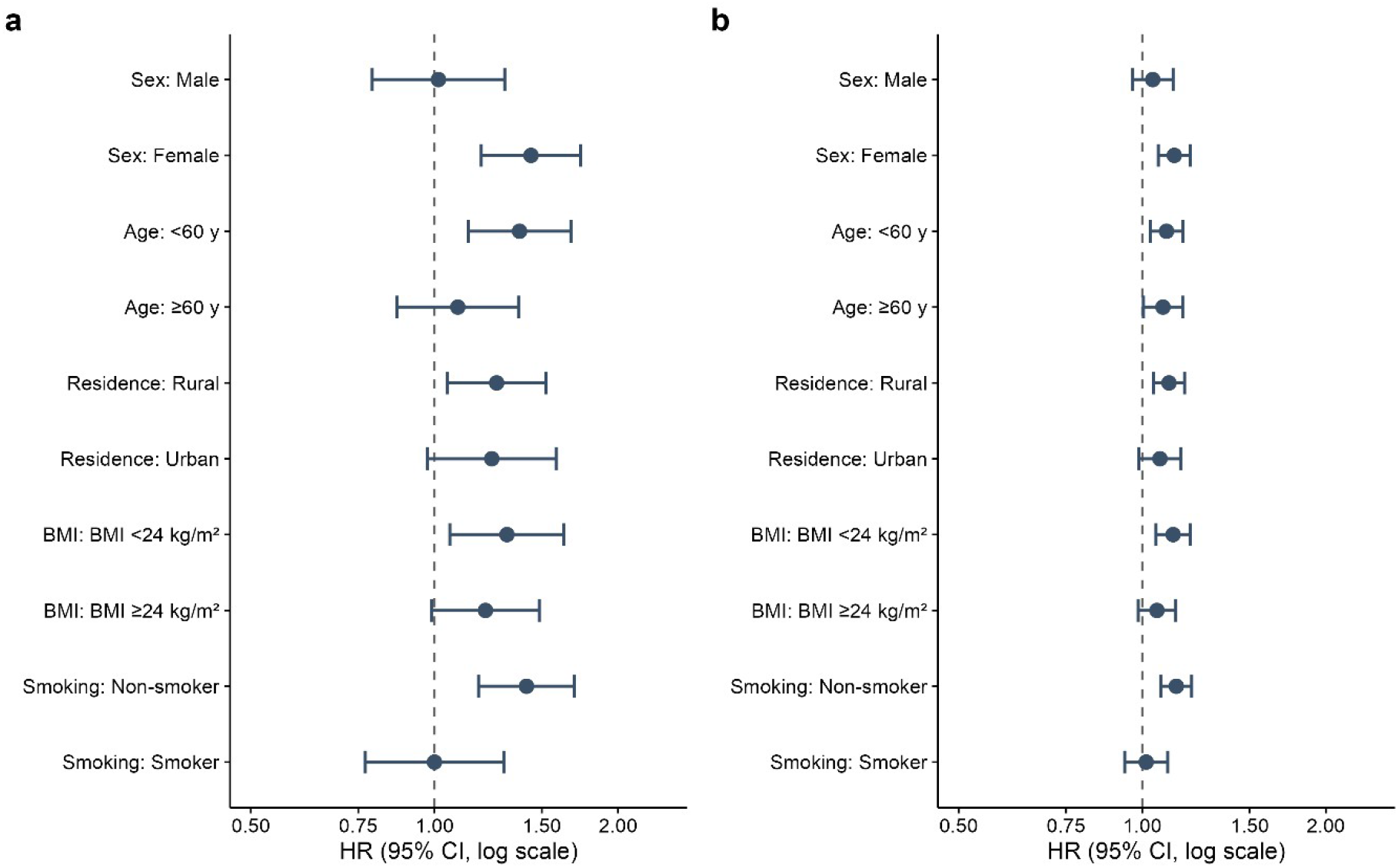
Forest plot of subgroup analyses. a, binary CircS (≥4 vs <4 components); b, continuous score (per one-component increase). Interaction P values were nominally significant only; none remained significant after Bonferroni correction.

### Sensitivity and negative-control analyses

The results remained robust after excluding events within 24 months of follow-up (HRs 1·107 and 1·283), excluding participants with baseline cardiovascular disease (1·082 and 1·252), restricting BMI to 18·5–35 kg/m^2^ (continuous and binary HRs 1·096 and 1·293), and using a CircS cutoff of ≥3 components (HR 1·280, 1·104–1·485) (Table S6). In Fine–Gray competing-risk regression, the subdistribution HR was 1·083 (1·034–1·134) per component and 1·235 (1·071–1·424) for CircS positivity. Under multiple imputation, estimates were slightly attenuated but remained significant (per one-component increase HR 1·081, 1·037– 1·128; CircS positive 1·223, 1·073–1·395). In the negative-control analysis, none of the four randomly generated exposures was associated with the outcome (HRs 0·99–1·12, all 95% CIs crossing 1; Table S5), suggesting no substantial residual confounding.

### Mediation by systemic inflammation

Baseline hs-CRP was positively associated with CircS (7·9% higher per additional component, P<0·001) but not with the outcome (b=−0·045, P=0·28); the bootstrap 95% CI of the indirect effect (a×b=−0·0034) included 0 (−0·011 to 0·003), with a mediated proportion of about −3·8%, indicating that systemic inflammation does not mediate the association (Figure S2).

## Discussion

Over 7 years of follow-up in this nationwide prospective cohort of middle-aged and older Chinese adults, CircS was independently associated with incident non-neoplastic digestive disease, with a 25·7% higher risk. Risk increased approximately linearly with the number of components, and participants with 5–7 components had an approximately 70% higher risk than those with none. The association was mainly driven by short sleep and depressive symptoms; none of the five metabolic components was independently associated in the mutually adjusted model. Although the association did not differ significantly between the first 36 months and thereafter (interaction P>0·05), baseline CircS remained associated with events occurring 4–7 years later, suggesting that the effect of CircS may take years to become apparent.

Our findings are largely consistent with a recent UK Biobank analysis of 330 925 adults, which also identified depression and short sleep as the strongest components of CircS for digestive outcomes^17^. Despite this agreement, the two studies differ in several aspects. First, the UK Biobank modelled the seven components separately, and each was associated with incident GERD. We fitted a mutually adjusted model containing all seven components, and only short sleep and depressive symptoms retained independent associations. This difference is expected, because the seven components overlap. When all are entered together, each component is adjusted for the others, so only independent associations remain. Second, the UK Biobank reported an HR of 1·20 for GERD in participants with versus without CircS, whereas our corresponding estimate was 1·257. The two estimates are close, but the outcomes differ, so they should not be compared directly. UK Biobank examined GERD only, using self-report and hospital records. Our study combined all self-reported, physician-diagnosed non-neoplastic digestive diseases. A combined outcome can dilute the association if CircS is more strongly related to some digestive diseases than to others. Third, the populations differ. UK Biobank participants were younger and healthier than the UK general population. CHARLS is broadly representative of Chinese adults aged 45 years or older, including rural residents and people with multiple chronic conditions. Differences in the underlying risk distribution between the two cohorts can shift relative estimates in either direction. Therefore, the small difference between the two HRs should not be overinterpreted. Fourth, modelling differed. The UK Biobank main models did not adjust for BMI, whereas our primary model did. In our data, BMI acted as a negative confounder: without BMI adjustment, the association was attenuated and no longer significant. Despite these differences, both studies point to the circadian rhythm–emotion axis as the dominant signal linking CircS to digestive disease.

Our finding that sleep and depression are the main components is consistent with previous studies of these factors individually. Prospective cohorts have linked short sleep to GERD and IBS^5,6^, and Mendelian randomization studies suggest a possible causal association between insomnia and peptic ulcer^22^. Depression is clearly associated with functional gastrointestinal disorders. The two conditions influence each other through the gut-brain axis^7,23^, and Mendelian randomization studies further suggest that this relationship may be causal^24^. Mechanistically, sleep deprivation and depression also share several pathophysiological processes that underlie the gut-brain axis. These include autonomic imbalance, increased hypothalamic-pituitary-adrenal axis activity, altered gastrointestinal motility, visceral sensitivity, and disruption of the gut microbiota and mucosal barrier^3,4,25^. These processes are closely linked to the enteric nervous system, so they can affect gastrointestinal motility, secretion, and visceral sensation through short, direct routes. Notably, systemic inflammation assessed by hs-CRP did not mediate the association, suggesting that the link is mediated more directly through the gut-brain axis than through systemic inflammation.

By contrast with these short, direct neural routes, obesity, dyslipidaemia, hypertension, and dysglycaemia affect the digestive system mainly through systemic low-grade inflammation and vascular mechanisms. These pathways are slower, less specific to the gut, and depend on exposure accumulated over many years. A possible explanation is that metabolic factors may help identify people with risk, but they do not independently increase risk. This explanation is consistent with previous evidence. Although several meta-analyses have linked metabolic syndrome to GERD and related oesophageal conditions^26,27^, the included studies did not adjust for sleep and mood, so their reported metabolic associations may partly reflect these co-occurring factors. The BMI analysis also illustrates how adjustment can change the apparent association. We retained BMI in the primary model for this reason and also reported estimates without BMI for comparison. Overall, previous findings on individual CircS components can be interpreted within the CircS framework. Metabolic components may be more useful for risk stratification than for mechanistic explanation.

If these findings are confirmed, prevention strategies for digestive diseases should not be limited to metabolic management. Sleep hygiene and emotional screening are low-cost, scalable interventions, and they are particularly suitable for individuals with multiple co-occurring circadian and mood disturbances. For sleep, cognitive behavioural therapy for insomnia (CBT-I) is the first-line non-pharmacological treatment for chronic insomnia. It has improved gastrointestinal symptoms in patients with functional gastrointestinal disorders in randomised trials, and digital delivery makes it feasible at the community level^28^. Brief structured psychological interventions, such as problem-solving therapy and behavioural activation, can be delivered by trained primary-care or community health workers^29^. These approaches have shown promise for depressive symptoms in primary-care settings. The higher point estimates in women, younger participants, and non-smokers, although non-significant after multiple-comparison correction and presented descriptively only, may provide preliminary clues for selecting target populations in intervention studies. In addition, risk increased with the number of components and was significantly elevated at 5–7 components, suggesting that the CircS score may serve as a practical tool for identifying high-risk individuals.

These inferences are supported by several design strengths but must be weighed against the study’s limitations. Strengths include the prospective design, the nationally representative sample, and causally informed covariate selection. Results were consistent across competing-risk, negative-control, and multiple-imputation sensitivity analyses. Several limitations should also be considered. First, the outcome was self-reported and aggregated heterogeneous diseases, so misclassification is possible. Such misclassification is generally non-differential and tends to underestimate the association. Event times were assigned by interview wave, resulting in interval censoring. The time-stratified results should therefore be regarded as hypothesis-generating only. Second, sleep and depression were both self-reported. The CES-D-10 contains somatic items that overlap with gastrointestinal symptoms, so common method bias remains possible. Sleep was assessed by self-reported duration only, without timing, quality, or regularity. Third, CircS was measured only once at baseline, and exposure misclassification is inevitable, tending to attenuate the estimates. In the mediation analysis, hs-CRP and the exposure were measured concurrently, so the results should be regarded as mechanistic exploration only. Fourth, the complete-case analysis retained 76·4% of participants with fasting blood samples. Excluded participants were older, more often women, and less educated, but their measured metabolic indicators were comparable. The multiple-imputation analysis yielded consistent conclusions. Finally, residual confounding by unmeasured factors such as diet and non-steroidal anti-inflammatory drug use cannot be ruled out, and our findings await confirmation in other populations.

## Conclusion

Among middle-aged and older Chinese adults, CircS is independently associated with incident non-neoplastic digestive disease in a dose–response manner. The association is mainly driven by short sleep and depressive symptoms, with no independent association for the metabolic components. Together with the UK Biobank study of GERD, our results suggest that the gut-brain axis may be a pathway linking modern lifestyles to digestive health, and that sleep and mood deserve attention in digestive disease prevention. Validation is needed in cohorts with objective sleep measurements, clinical gastrointestinal endpoints, and repeated exposure assessments.

## Supporting information

STROBE checklist

## Data Availability

Deidentified CHARLS data are available from the CHARLS project team upon application (https://charls.pku.edu.cn), subject to approval of a research proposal and a signed data use agreement. The statistical analysis code is available from the corresponding author upon reasonable request. No other datasets were used.

## Declarations

### Ethics approval and consent to participate

the study was approved by the Biomedical Ethics Review Committee of Peking University (approval number: IRB00001052-11015 for household survey and IRB00001052-11014 for blood sample), and all participants provided written informed consent.

### Conflicts of interest

The authors declare no competing interests.

### Contributors

Lu Liu and Liqiong Zhao contributed equally to this work and are designated as co-first authors. Lu Liu, Liqiong Zhao, and Yan Geng designed the study. Lu Liu and Cailan Li performed the statistical analysis. Liqiong Zhao had full access to all the data in the study and verified the underlying data. Lu Liu and Cailan Li drafted the manuscript. All authors critically revised the manuscript and approved the final version. Yan Geng supervised the study and is the guarantor.

### Data sharing

Deidentified CHARLS participant data and the data dictionary will be available with publication of this Article for 10 years, from the CHARLS project team upon application (https://charls.pku.edu.cn), subject to approval of a research proposal and a signed data use agreement. The study protocol and informed consent forms are available from the same source. The statistical analysis plan and analysis code are available from the corresponding author on request, subject to the same access criteria; the code requires access to the CHARLS core, health-care, and blood examination datasets.

### Declaration of generative AI use

Generative AI tools (Kimi and DeepSeek) were used to assist with language polishing and formatting. All scientific content, hypotheses, data analysis, interpretation, and conclusions were independently developed by the authors.

## Acknowledgments

We thank the CHARLS research team and all participants.

## Funding

This research received no specific grant from any funding agency in the public, commercial, or not-for-profit sectors.

## Notes

### Competing Interest Statement

The authors have declared no competing interest.

### Author Declarations

Biomedical Ethics Review Committee of Peking University gave ethical approval for this work (approval numbers IRB00001052-11015 for the household survey and IRB00001052-11014 for blood sampling).

