## Supplementary appendix for "Circadian syndrome and incident non-neoplastic digestive disease in middle-aged and older Chinese adults: a prospective cohort study"

This file contains Figures S1–S2b and Tables S1–S9, numbered as cited in the main text.

### Contents

Figure S1. Event-free survival curves by CircS component group after the 36-month landmark
Figure S2. hs-CRP mediation analysis path diagram (continuous exposure)
Figure S2b. hs-CRP mediation analysis path diagram (binary exposure)
Table S1. Comparison of included and excluded participants
Table S2. Period-by-exposure interaction tests (piecewise Cox models, LRT)
Table S3. Time-stratified analysis
Table S4. Component-specific associations
Table S5. Negative-control analysis
Table S6. Sensitivity analyses
Table S7. Schoenfeld proportional hazards assumption test
Table S8. Association between the number of CircS components and the outcome
Table S9. Subgroup analyses

**Figure S1.** Event-free survival curves by CircS component group (0–1, 2–3, 4–7 components) after the 36-month landmark (log-rank P=0·072).

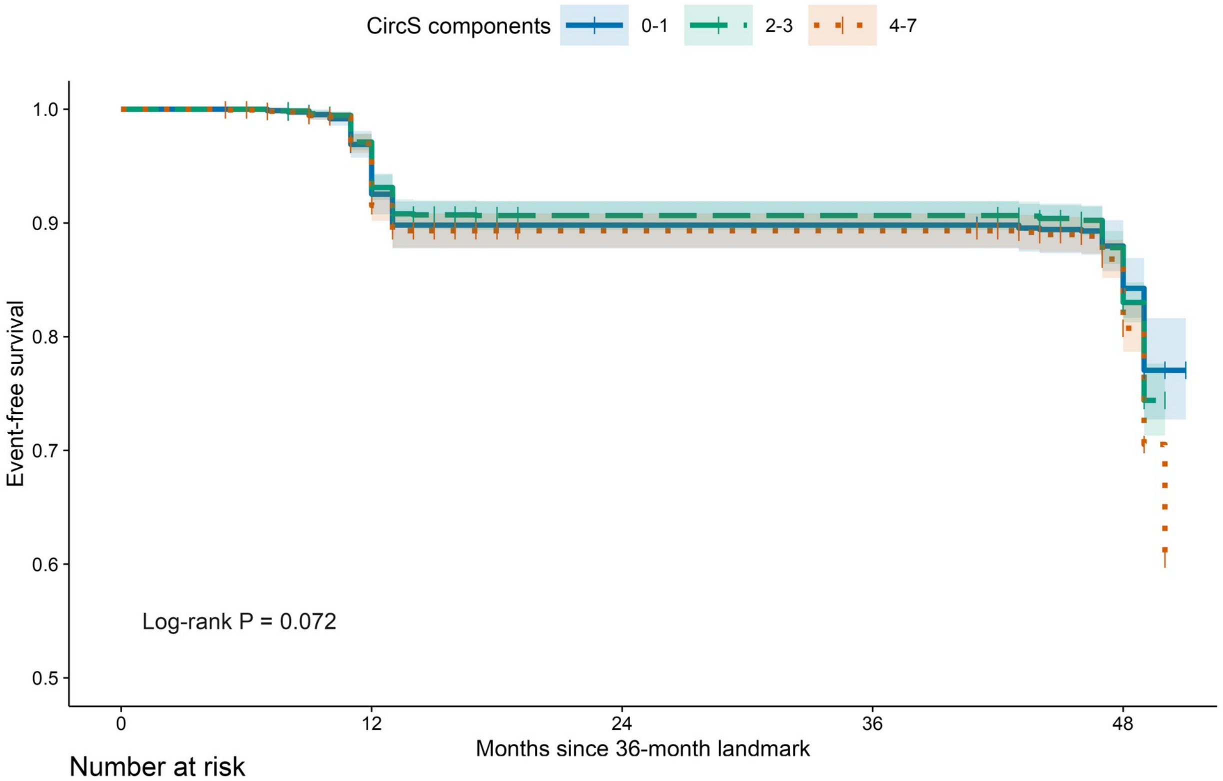


Number at risk

| | 0 | 12 | 24 | 36 | 48 |
|---|---|---|---|---|---|
| 0-1 | 843 | 798 | 706 | 706 | 520 |
| 2-3 | 2055 | 1967 | 1724 | 1724 | 1256 |
| 4-7 | 1632 | 1565 | 1335 | 1335 | 957 |

**Figure S2.** hs-CRP mediation analysis path diagram (continuous exposure, per +1 component). a: exposure→mediator path (linear regression coefficient); b: mediator→outcome path (Cox model log-HR); c′: direct effect. Indirect effect a×b=−0·0034 (1 000 bootstrap resamples, 95% CI −0·0105 to 0·0032, including 0); mediated proportion −3·8%. Mediation is not supported.

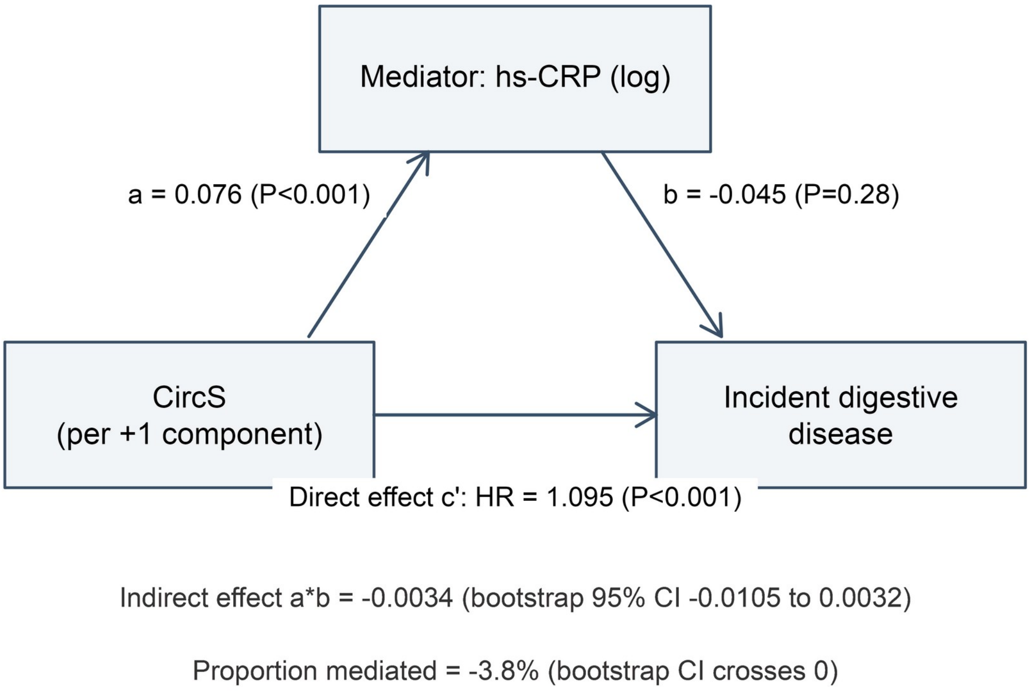

**Figure S2b.** hs-CRP mediation analysis path diagram (binary exposure, ≥4 vs <4 components). a=0·211 (P<0·001); b=−0·040 (P=0·33); direct effect HR=1·268 (P=0·002); indirect effect a×b=−0·0085 (95% CI −0·0274 to 0·0093); mediated proportion −3·8%.

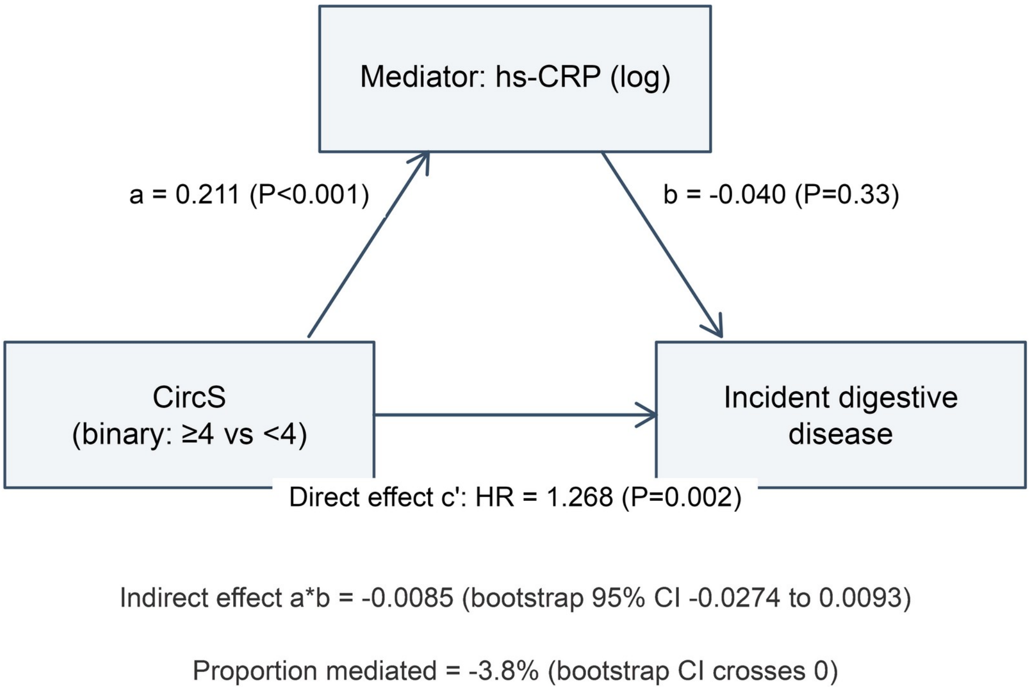

**Table S1. Comparison of included and excluded participants (6 243 fasting-eligible participants)**

| Characteristic | Overall (n=6 243) | Included (n=4 771) | Excluded (n=1 472) | P | SMD |
|---|---|---|---|---|---|
| Age, years | 59·6±9·4 | 58·9±8·9 | 62·0±10·4 | <0·001 | 0·327 |
| Female, n (%) | 3 271 (52·4) | 2 313 (48·5) | 958 (65·1) | <0·001 | 0·340 |
| BMI, kg/m² | 23·6±3·7 | 23·8±3·6 | 23·2±3·8 | <0·001 | 0·159 |
| Education: senior high school or above | 653 (10·5) | 589 (12·3) | 64 (4·4) | <0·001 | 0·471 |
| Education: junior high school | 1 268 (20·3) | 1 102 (23·1) | 166 (11·3) | | |
| Education: primary school or below | 4 320 (69·2) | 3 080 (64·6) | 1 240 (84·4) | | |
| Married, n (%) | 5 428 (86·9) | 4 248 (89·0) | 1 180 (80·2) | <0·001 | 0·248 |
| Rural residence, n (%) | 3 938 (63·1) | 2 894 (60·7) | 1 044 (70·9) | <0·001 | 0·218 |
| Smoking, n (%) | 2 457 (39·4) | 2 000 (41·9) | 457 (31·1) | <0·001 | 0·227 |
| Alcohol use, n (%) | 2 619 (42·0) | 2 118 (44·4) | 501 (34·2) | <0·001 | 0·211 |
| Triglycerides, mg/dL, median [IQR] | 105·3 [75·2, 152·2] | 107·1 [75·2, 154·9] | 100·9 [73·5, 145·1] | <0·001 | 0·113 |
| HDL-C, mg/dL | 51·2±15·3 | 50·8±15·2 | 52·6±15·6 | <0·001 | 0·120 |
| Fasting glucose, mg/dL, median [IQR] | 102·4 [94·7, 112·9] | 102·4 [94·9, 112·9] | 102·1 [94·3, 112·7] | 0·457 | 0·010 |
| Systolic blood pressure, mmHg | 130·2±21·3 | 129·8±20·8 | 131·6±22·6 | 0·004 | 0·084 |
| Diastolic blood pressure, mmHg | 75·8±12·3 | 76·0±12·2 | 75·1±12·5 | 0·029 | 0·066 |

Note: excluded participants were older, more often women, and less educated; however, measured metabolic profiles (fasting glucose, triglycerides, HDL-C, blood pressure) were highly similar between the two groups (all SMDs <0·13), suggesting limited selection bias at the exposure assessment level.

**Table S2. Period-by-exposure interaction tests (piecewise Cox models, LRT)**

| Exposure | Interaction HR (95% CI) | LRT P |
|---|---|---|
| CircS (continuous) | 1·086 (0·964–1·225) | 0·174 |
| CircS (binary) | 1·108 (0·749–1·641) | 0·607 |

**Table S3. Time-stratified analysis (secondary; model 3 covariates)**

| Period | Continuous HR (95% CI) | Binary HR (95% CI) | Events |
|---|---|---|---|
| 0–36 months | 0·996 (0·876–1·132), P=0·95 | 1·127 (0·753–1·687), P=0·56 | 123 |
| >36 months (landmark) | 1·107 (1·053–1·165), P<0·001 | 1·279 (1·092–1·499), P=0·002 | 784 |

**Table S4. Component-specific associations (model 3 covariates)**

| Component | Mutually adjusted HR (95% CI), P | Single-component HR (95% CI), P |
|---|---|---|
| Impaired fasting glucose | 1·028 (0·897–1·179), 0·69 | 1·030 (0·900–1·178), 0·67 |
| Elevated triglycerides | 1·051 (0·893–1·236), 0·55 | 1·033 (0·889–1·201), 0·67 |
| Reduced HDL-C | 0·949 (0·813–1·107), 0·50 | 0·971 (0·841–1·122), 0·69 |
| Elevated blood pressure | 0·950 (0·827–1·092), 0·47 | 0·966 (0·841–1·109), 0·62 |
| Central obesity | 1·074 (0·902–1·279), 0·42 | 1·085 (0·912–1·289), 0·36 |
| Short sleep | 1·178 (1·017–1·365), 0·029 | 1·291 (1·117–1·491), <0·001 |
| Depressive symptoms | 1·570 (1·367–1·803), <0·001 | 1·617 (1·411–1·852), <0·001 |

**Table S5. Negative-control analysis (random exposures, model 3 covariates)**

| Random exposure | HR (95% CI) | P |
|---|---|---|
| Normal continuous | 1·026 (0·961–1·095) | 0·44 |
| Uniform continuous | 1·115 (0·893–1·394) | 0·34 |
| Binary (p=0·5) | 0·994 (0·873–1·133) | 0·93 |
| Binary (p=0·3) | 1·031 (0·896–1·187) | 0·67 |

**Table S6. Sensitivity analyses (model 3 covariates unless otherwise noted)**

| Analysis | Continuous HR (95% CI) | Binary HR (95% CI) | n/events |
|---|---|---|---|
| Primary analysis (≥4 cutoff) | 1·091 (1·041–1·144) | 1·257 (1·084–1·456) | 4 771/907 |
| Alternative cutoff ≥3 | 1·091 (1·041–1·144) | 1·280 (1·104–1·485) | 4 771/907 |
| Without BMI | 1·034 (0·991–1·079) | 1·106 (0·965–1·269) | 4 771/907 |
| BMI restricted to 18·5–35 | 1·096 (1·043–1·151) | 1·293 (1·111–1·505) | 4 517/832 |
| Excluding baseline CVD | 1·082 (1·028–1·139) | 1·252 (1·066–1·470) | 4 194/782 |
| Excluding events within 24 months | 1·107 (1·054–1·163) | 1·283 (1·099–1·497) | 4 688/824 |
| Multiple imputation (20 sets) | 1·081 (1·037–1·128) | 1·223 (1·073–1·395) | ≈6 140/— |
| Fine–Gray (death as competing event) | 1·083 (1·034–1·134) | 1·235 (1·071–1·424) | 4 771/907 |

**Table S7. Schoenfeld proportional hazards assumption test (model 3; cox.zph, transform="km")**

| Variable | $\chi^2$ | df | P |
|---|---|---|---|
| CircS component score (continuous) | 3·975 | 1 | 0·046 |
| Age | 3·818 | 1 | 0·051 |
| Sex | 0·072 | 1 | 0·788 |
| Education | 0·950 | 2 | 0·622 |
| Marital status | 0·619 | 1 | 0·431 |
| Residence | 2·073 | 1 | 0·150 |
| BMI | 3·104 | 1 | 0·078 |
| Smoking | 0·074 | 1 | 0·786 |
| Alcohol use | 0·656 | 1 | 0·418 |
| Global test | 13·222 | 10 | 0·212 |

Note: the global test was not significant (P=0·212) and no covariate showed a violation (all P≥0·05); only CircS showed borderline time variation (P=0·046), on which basis the time-stratified analyses (Tables S2 and S3) were performed, as hypothesis-generating only.

**Table S8. Association between the number of CircS components and incident digestive disease (model 3 covariates)**

| Number of CircS components | HR (95% CI) |
|---|---|
| 0 (reference) | 1·00 |
| 1 | 1·143 (0·800–1·633) |
| 2 | 1·149 (0·816–1·618) |
| 3 | 1·355 (0·961–1·911) |
| 4 | 1·422 (0·997–2·029) |
| 5–7 | 1·698 (1·182–2·440) |

Note: P for linear trend <0·001; the dose–response curve is shown in Figure 2 (non-linearity P=0·76).

**Table S9. Subgroup analyses (model 3 covariates)**

| Subgroup | n | Events | Continuous (per +1 component) HR (95% CI), P | Binary (≥4 vs <4) HR (95% CI), P |
|---|---|---|---|---|
| Sex: men | 2 458 | 391 | 1·040 (0·963–1·123), 0·313 | 1·015 (0·790–1·304), 0·907 |
| Sex: women | 2 313 | 516 | 1·129 (1·062–1·199), <0·001 | 1·440 (1·193–1·738), <0·001 |
| Interaction P | | | 0·106 | 0·032 |
| Age: <60 years | 2 683 | 522 | 1·096 (1·030–1·166), 0·004 | 1·379 (1·136–1·674), 0·001 |
| Age: ≥60 years | 2 088 | 385 | 1·081 (1·004–1·164), 0·038 | 1·092 (0·868–1·374), 0·452 |
| Interaction P | | | 0·335 | 0·049 |
| Residence: rural | 2 894 | 585 | 1·106 (1·043–1·173), <0·001 | 1·265 (1·049–1·524), 0·014 |
| Residence: urban | 1 877 | 322 | 1·069 (0·988–1·157), 0·099 | 1·241 (0·973–1·583), 0·082 |
| Interaction P | | | 0·984 | 0·648 |
| BMI: <24 kg/m² | 2 682 | 509 | 1·124 (1·053–1·199), <0·001 | 1·315 (1·060–1·631), 0·013 |
| BMI: ≥24 kg/m² | 2 089 | 398 | 1·057 (0·985–1·134), 0·123 | 1·212 (0·989–1·486), 0·063 |
| Interaction P | | | 0·398 | 0·806 |
| Smoking: no | 2 771 | 570 | 1·137 (1·073–1·205), <0·001 | 1·416 (1·181–1·697), <0·001 |
| Smoking: yes | 2 000 | 337 | 1·015 (0·936–1·101), 0·720 | 1·000 (0·770–1·300), 1·000 |
| Interaction P | | | 0·134 | 0·122 |

Note: 10 interaction tests in total; none remained significant after Bonferroni correction (α=0·005), and subgroup differences are presented descriptively only. Corresponds to Figure 4.
