## Supplementary material for "Circadian syndrome and incident non-neoplastic digestive disease in middle-aged and older Chinese adults: a prospective cohort study": STROBE checklist

**STROBE Statement—Checklist of items for cohort studies**

Manuscript: "Circadian syndrome and incident non-neoplastic digestive disease in middle-aged and older Chinese adults: a prospective cohort study". Page and line numbers refer to the submitted manuscript file (manuscript.docx, 22 pages), which includes continuous line numbering.

| **Item** | **Recommendation** | **Location in manuscript** |
| --- | --- | --- |
| 1a Title and abstract | Indicate the study's design with a commonly used term in the title or the abstract | Title page (p1, lines 1–3); Abstract (p2, lines 28–52) |
| 1b Title and abstract | Provide in the abstract an informative and balanced summary of what was done and what was found | Abstract (p2, lines 28–52) |
| 2 Introduction | Scientific background and rationale | Introduction (pp3–4, lines 57–89) |
| 3 Introduction | Specific objectives or hypotheses | Introduction (p4, lines 90–96) |
| 4a Methods: Study design | Present key elements of study design early in the paper | Methods—Study design and population (p4, lines 97–110) |
| 4b Methods: Setting | Setting, locations, relevant dates | Methods—Study design and population (p4, lines 97–104) |
| 4c Methods: Participants | Eligibility criteria; sources and methods of selection | Methods—Study design and population (p4, lines 105–110) |
| 4d Methods: Variables | Outcomes, exposures, predictors, confounders | Methods—Exposure (p4, lines 112–125); Outcome (p5, lines 126–135); Covariates (p5, lines 136–151) |
| 4e Methods: Data sources/measurement | Assessment of each variable | Methods—Exposure; Outcome; Covariates (pp4–5, lines 112–151) |
| 4f Methods: Bias | Efforts to address potential sources of bias | Methods—Covariates (p5, lines 146–147); Results—baseline characteristics (p8, lines 201–205; Table S1); Discussion—limitations (p13, lines 351–371) |
| 4g Methods: Study size | How study size was arrived at | Methods—Study design and population (p4, lines 105–110) |
| 4h Methods: Quantitative variables | How quantitative variables were handled in the analyses | Methods—Exposure (p4, lines 112–125); Statistical analysis (p6, lines 152–183) |
| 4i(a) Methods: Statistical methods | Describe all statistical methods, including those used to control for confounding | Methods—Statistical analysis (p6, lines 156–183) |
| 4i(b) Methods: Statistical methods | Describe any methods used to examine subgroups and interactions | Methods—Statistical analysis (p6, lines 174–178) |
| 4i(c) Methods: Statistical methods | Explain how missing data were addressed | Methods—Statistical analysis (p6, lines 182–183) |
| 4i(d) Methods: Statistical methods | Cohort study—if applicable, explain how loss to follow-up was addressed | Methods—Outcome (p5, lines 130–134) |
| 4i(e) Methods: Statistical methods | Describe any sensitivity analyses | Methods—Statistical analysis (pp6–7, lines 179–186) |
| 5a Results: Participants | Report numbers of individuals at each stage of the study | Methods—Study design and population (p4, lines 105–110); Figure 1 (p19, legend line 486) |
| 5b Results: Participants | Give reasons for non-participation at each stage | Figure 1 (p19, legend line 486); Table S1 (supplementary appendix, p5); Results—baseline characteristics (p8, lines 201–205) |
| 5c Results: Participants | Consider use of a flow diagram | Figure 1 (p19, legend line 486) |
| 6a Results: Descriptive data | Give characteristics of study participants and information on exposures and potential confounders | Results—Baseline characteristics (p8, lines 195–205); Table 1 (p17) |
| 6b Results: Descriptive data | Indicate the number of participants with missing data for each variable of interest | Methods—Statistical analysis (p6, lines 182–183); Figure 1 (p19) |
| 7 Results: Outcome data | Report numbers of outcome events or summary measures | Results—Baseline characteristics (p8, lines 197–198) |
| 8a Results: Main results | Give unadjusted estimates and, if applicable, confounder-adjusted estimates and their precision | Results—CircS and risk of digestive disease (p8, lines 206–221); Table 2 (p18) |
| 8b Results: Main results | Report category boundaries when continuous variables were categorised | Methods—Exposure (p5, lines 122–125); Table S8 (supplementary appendix, p12) |
| 8c Results: Main results | If relevant, consider translating estimates of relative risk into absolute risk for a meaningful time period | Results—Dose–response relationship (p8, lines 222–228); Table S8; Figure 2 (p20, legend line 488) |
| 9 Results: Other analyses | Report other analyses done—eg analyses of subgroups and interactions, and sensitivity analyses | Results—Component-specific associations (p9, lines 239–245); Subgroup analysis (p9, lines 246–252); Sensitivity and negative-control analyses (p9, lines 253–263); Mediation (p10, lines 264–269); Tables S2–S7 |
| 10a Discussion | Summarise key results with reference to study objectives | Discussion (p11, lines 272–281) |
| 10b Discussion | Discuss limitations of the study, taking into account sources of bias and imprecision | Discussion—limitations (p13, lines 351–371) |
| 10c Discussion | Give a cautious overall interpretation of results considering objectives, limitations, multiplicity of analyses, results from similar studies, and other relevant evidence | Discussion (pp11–13, lines 282–371) |
| 10d Discussion | Discuss the generalisability (external validity) of the study results | Discussion (p13, lines 351–371) |
| 11 Other information: Funding | Give the source of funding and the role of the funders for the present study | Declarations—Funding (p15, line 405); Abstract (p2, lines 49–50) |
